# Rare but Relevant: Assessing Variants in Dystonia-linked Genes in Parkinson’s Disease

**DOI:** 10.1101/2025.07.10.25330831

**Authors:** Lara M. Lange, Zih-Hua Fang, Laurel Screven, Ai Huey Tan, Roy N. Alcalay, Rim Amouri, Roberta Bovenzi, Matilda Fenn, Joshua L. I. Frost, Joseph Jankovic, Simona Jasaityte, Zane Jaunmuktane, Beomseok Jeon, Ignacio Juan Keller Sarmiento, Rejko Krüger, Gregor Kuhlenbäumer, Chin-Hsien Lin, Lukas Pavelka, Maria Teresa Periñan, Samia Ben Sassi, Tommaso Schirinzi, Jung Hwan Shin, Joshua M. Shulman, Yi Wen Tay, Ryan Uitti, Tom Warner, Zbigniew K. Wszolek, Lesley Wu, Ruey-Meei Wu, Kirsten E. Zeuner, Cornelis Blauwendraat, Andrew Singleton, Niccolò E. Mencacci, Huw R. Morris, Shen-Yang Lim, Katja Lohmann, Christine Klein, Global Parkinson’s Genetics Program (GP2)

## Abstract

**Background:** Dystonia and Parkinson’s disease (PD) show clinical and genetic overlap, but the relevance of dystonia gene variants in PD remains unclear.

**Objective:** To assess the frequency of dystonia-linked pathogenic variants in PD.

**Methods:** We screened sequencing data from 15,738 individuals (7,851 PD, 4,287 atypical parkinsonism, and 3,600 unaffected) from GP2 and AMP-PD for variants in genes linked to isolated dystonia, dystonia-parkinsonism, and myoclonus-dystonia.

**Results:** Pathogenic variants were only identified in PD patients. Forty-five PD individuals (0.57%) carried 26 distinct (likely) pathogenic variants in nine dystonia-linked genes, most frequently in *GCH1*, followed by *VPS16*.

**Conclusion:** Though rare, pathogenic variants in dystonia-linked genes are present in clinically and pathologically diagnosed PD. Our results reinforce *GCH1* as a PD-relevant gene with clinical implications, while variants identified in other genes are rare and of sometimes uncertain relation to the PD phenotype.

## Introduction

Dystonia is a clinically heterogeneous disorder. Its etiology includes nervous system pathologies, acquired causes, and genetic factors. (1) To date, variants in over 400 genes have been linked to different forms of dystonia, though the majority are associated with more complex and broader neurological presentations. (2–7) The term “dystonia” is not only used to refer to a disease entity itself but also to describe a symptom as part of another neurological disorder. For example, dystonic symptoms are frequently reported in individuals with Parkinson’s disease (PD), either related to dopaminergic treatment and motor fluctuations or as an initial disease manifestation, especially in early-onset PD. (8,9) Dystonic symptoms are also commonly encountered in atypical parkinsonism, though the dystonic features typically differ from those in PD. Interestingly, previous screening studies of PD patients identified carriers of pathogenic variants in genes primarily linked to dystonia, most frequently *GCH1*. (10,11)

Moreover, several neurogenetic conditions include features of both dystonia and parkinsonism, either individually or combined, i.e., where dystonia and parkinsonism are equally prominent. Based on this phenotypic and genetic overlap between dystonia and parkinsonism, this study aimed to assess the frequency of pathogenic variants in dystonia genes in individuals with PD and atypical parkinsonism by leveraging large-scale sequencing data from the Global Parkinson’s Genetics Program (GP2; https://gp2.org/) (12,13) and the Accelerating Medicines Partnership - Parkinson’s disease (AMP-PD; https://www.amp-pd.org/).

## Methods

Figure 1 illustrates our workflow. We analyzed short-read sequencing data from 15,738 individuals of eleven genetically determined ancestries from GP2’s Data Release 8 (DOI 10.5281/zenodo.13755496) and AMP-PD’s Release 4, including 7,851 PD patients, 4,287 individuals with atypical parkinsonism, and 3,600 unaffected individuals (see Supplementary Material).

**Figure 1.**
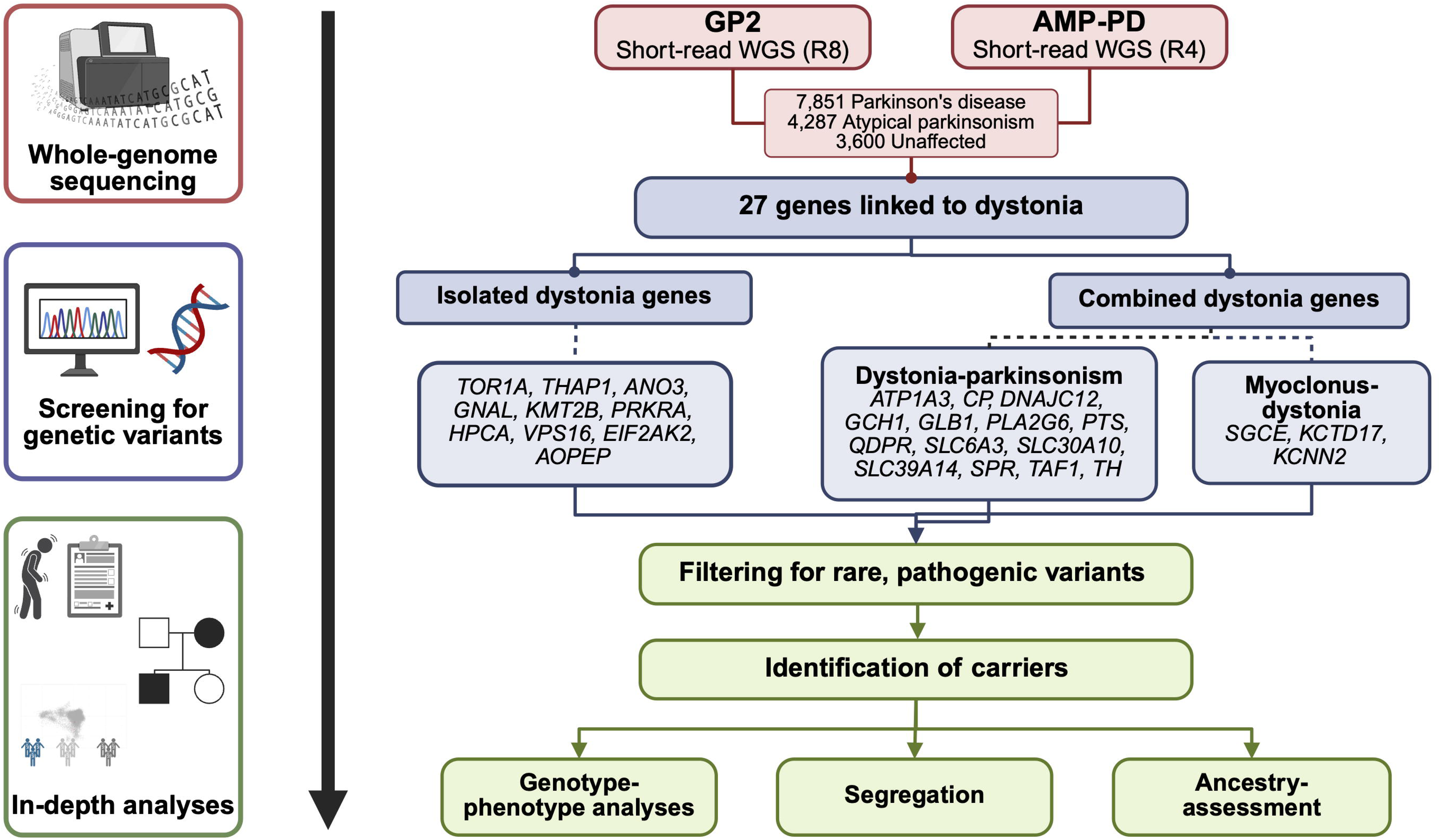
Study design and workflow. We screened short-read whole-genome sequencing (WGS) data from the Global Parkinson’s Genetics Program (GP2; Release 8 [R8]) and the Accelerating Medicines Partnership - Parkinson’s disease (AMP-PD, Release 4 [R4]) for known variants in genes linked to different forms of dystonia. We included genes linked to isolated as well as combined dystonia phenotypes, the latter including dystonia-parkinsonism and myoclonus-dystonia. Only rare variants predicted to be pathogenic or likely pathogenic were included in further analyses. For identified carriers, we evaluated genotype-phenotype correlations, investigated segregation, and assessed ancestry distributions, where possible. This figure was created with BioRender.

We investigated variants in genes linked to dystonia following the recommendations of the *MDS Task Force on the Nomenclature of Genetic Movement Disorders* (*2*): i) isolated dystonia: *ANO3, AOPEP, EIF2AK2, GNAL, HPCA, KMT2B, PRKRA, THAP1, TOR1A,* and *VPS16*, ii) dystonia-parkinsonism: *ATP1A3, CP, DNAJC12, GCH1, GLB1, PLA2G6, PTS, QDPR, SLC6A3, SLC30A10, SLC39A14, SPR, TAF1,* and *TH*, and iii) myoclonus-dystonia: *SGCE, KCTD17,* and *KCNN2*. We filtered for rare (gnomAD minor allele frequency ≤1%) variants classified as pathogenic/likely pathogenic according to ClinVar (https://www.ncbi.nlm.nih.gov/clinvar/) and ACMG criteria (14) (see Supplementary Material).

All code generated for this article, and the identifiers for all software programs and packages used, are available on GitHub (https://github.com/GP2code/dystonia-genes-inPD) and were given a persistent identifier via Zenodo (DOI 10.5281/zenodo.15676002).

## Results

We identified 45 individuals from six ancestries carrying 26 distinct pathogenic/likely pathogenic variants in nine dystonia-linked genes. The identified pathogenic/likely pathogenic variants and their pathogenicity evaluation are summarized in Table 1. All 45 individuals were diagnosed with PD (n=45/7,851; 0.57%), while no carriers were diagnosed with atypical parkinsonism (n=0/4,287; 0%) or among unaffected individuals (n=0/3,600; 0%). The majority (n=43) were carriers of heterozygous variants in genes linked to dominantly inherited dystonia genes (*ATP1A3, GCH1, SGCE, KMT2B, THAP1, TOR1A,* and *VPS16*), while only one carrier of a homozygous *PLA2G6* variant and one of compound-heterozygous *PTS* variants (Supplementary Figure 3) were identified.

**Table 1.**
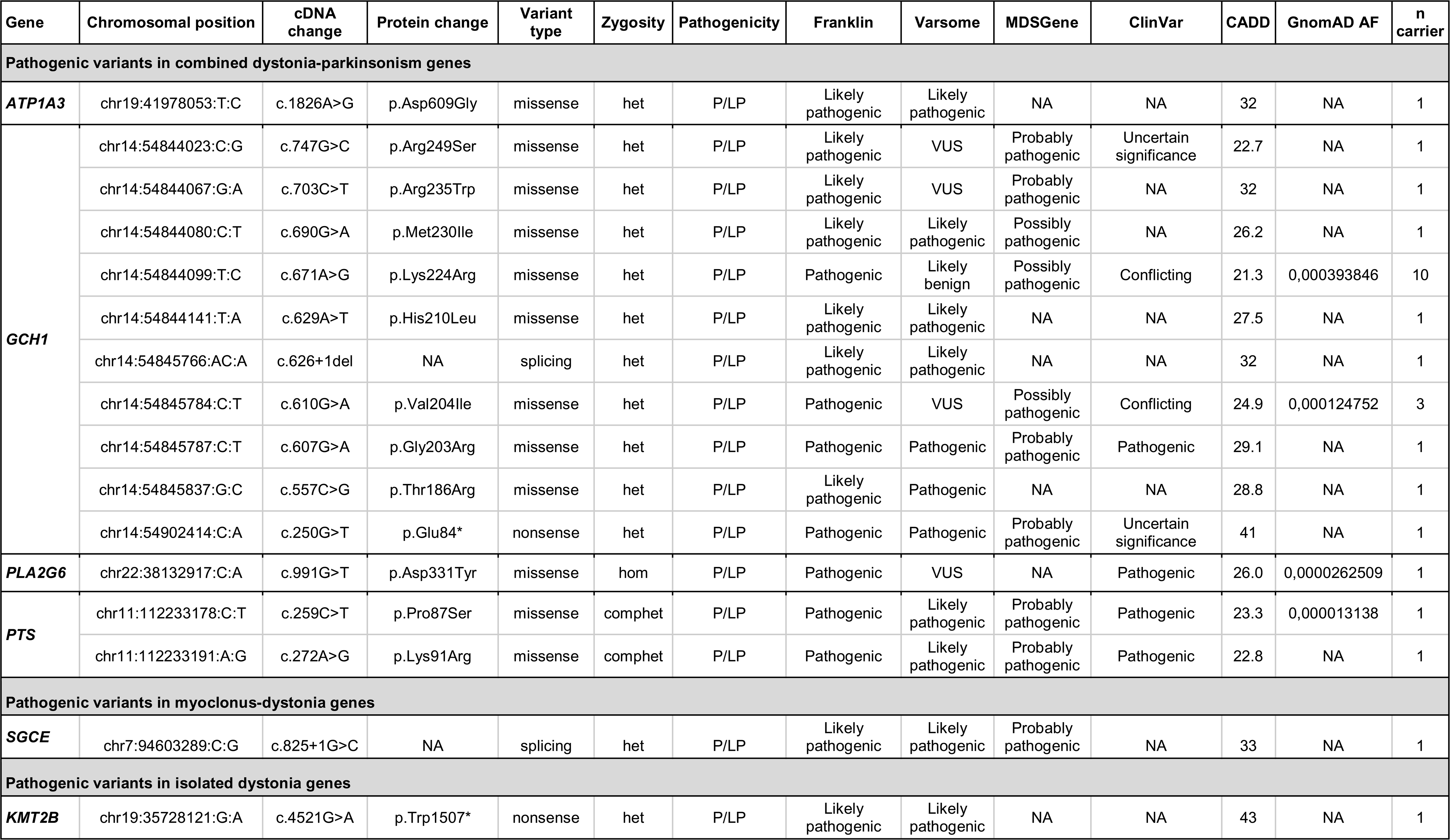
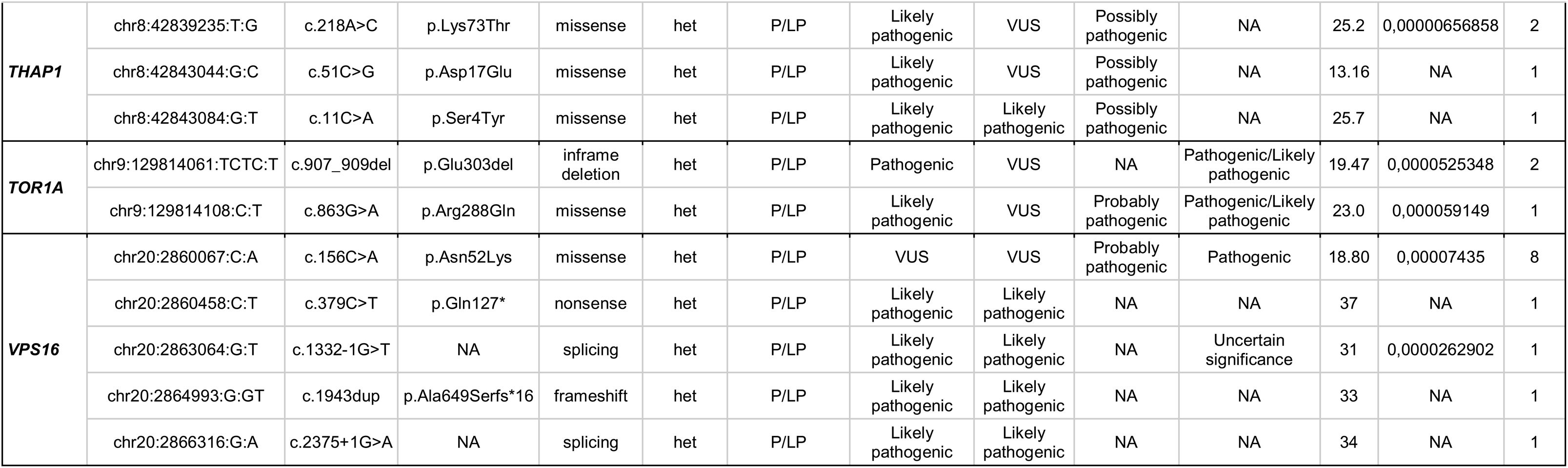
Overview of identified pathogenic and likely pathogenic variants in dystonia-linked genes. AF = allele frequency, comphet = compound heterozygous, het = heterozygous, hom = homozygous, NA = not available/not applicable, P/LP = pathogenic/likely pathogenic, VUS = variant of uncertain significance CADD: Combined Annotation Dependent Depletion; https://cadd.gs.washington.edu/; ClinVar: https://www.ncbi.nlm.nih.gov/clinvar/; GnomAD: https://gnomad.broadinstitute.org/ (version 4.1.0); Franklin: https://franklin.genoox.com; MDSGene: https://www.mdsgene.org/; Varsome: https://varsome.com/

Overall, variants in *GCH1* were the most frequent (n=21/45, 46.7%), followed by *VPS16* (n=12/45, 26.7%). Notably, there was one ancestry-specific recurrent variant in both these genes: *GCH1* p.Lys224Arg, carried by ten European-ancestry individuals, including two siblings, and *VPS16* p.Asn52Lys, carried by eight East Asian individuals. According to gnomAD, both variants are more frequent in these respective ancestral groups compared to other ancestries; however, the frequency in PD cases in our study was higher compared to ancestry-matched controls (Supplementary Table 2). With respect to additional variants in PD-linked genes, seven of the 45 individuals carried *GBA1* or *LRRK2* risk variants, and two carried heterozygous pathogenic variants in recessively inherited PD genes (Table 2), but none harbored a disease-explaining variant in an established PD-linked gene (including *LRRK2, SNCA, VPS35, RAB32, PINK1, PRKN,* and *PARK7/DJ-1*).

**Table 2.** Demographic and clinical characteristics of identified variant carriers. AAC = African admixed ancestry, AD = Alzheimer’s disease, AJ = Ashkenazi Jewish, ARTAG = Aging-Related Tau Astrogliopathy, CAS = Central Asian ancestry, CAA = Cerebral Amyloid Angiopathy, EAS = East Asian ancestry, EUR = European, FTLD = Frontotemporal Lobar Degeneration, het = heterozygous, LBD = Lewy Body Dementia, MDE = Middle Eastern ancestry, PD = Parkinson’s disease, RBD = REM sleep behavior disorder, SVD = Small Vessel Disease [1] When both were available, the AAO is displayed. For those individuals without an available AAO, we indicate the AAD. According to MedRxiv’s regulations, AAO/AAD is provided in ranges. * The asterisk highlights related individuals within the investigated cohort. GP2-ID-5 and GP2-ID-6 are siblings, and GP2-ID-20 is the daughter of GP2-ID-21. # This symbol highlights individuals who underwent deep brain stimulation (DBS).

| ID | Gender | Genetic ancestry | Gene | Variant | Additional genetic finding | Diagnosis | AAO/ AAD [1] | Symptom at onset | Levodopa responsive | FH PD | FH Dystonia | Details FH | Signs of Dystonia | Atypical signs |
| --- | --- | --- | --- | --- | --- | --- | --- | --- | --- | --- | --- | --- | --- | --- |
| Pathogenic variants in combined dystonia-parkinsonism genes |  |  |  |  |  |  |  |  |  |  |  |  |  |  |
| GP2-ID-1 | Male | EUR | *ATP1A3* | p.Asp609Gly |  | PD | 26-30 | NA | Yes | No | No | NA | Severe OFF dystonia | NA |
| GP2-ID-2 | Male | EAS | *GCH1* | p.Arg249Ser |  | PD | 66-70 | Tremor and slowness | Yes | Yes | No | Brother with PD | No | No; dementia after 10 years disease duration |
| GP2-ID-3 | Female | AAC | *GCH1* | p.Arg235Trp |  | PD | 31-35 | Gait disorder | Yes | No | No | NA | Cervical dystonia | Mental retardation |
| GP2-ID-4 | Female | CAS | *GCH1* | p.Met230Ile |  | PD | 41-45 | NA | NA | No | NA | NA | NA | NA |
| GP2-ID-5* | Female | EUR | *GCH1* | p.Lys224Arg |  | PD | 71-75 | NA | NA | Yes | NA | Sister with PD | NA | No |
| GP2-ID-6* | Female | EUR | *GCH1* | p.Lys224Arg |  | PD | 56-60 | NA | NA | Yes | NA | Sister with PD | NA | No |
| GP2-ID-7 | Male | EUR | *GCH1* | p.Lys224Arg |  | PD | 36-40 | NA | NA | Yes | NA | Father with possible PD (not diagnosed) | NA | NA |
| AMPP D-ID-1 | Female | EUR | *GCH1* | p.Lys224Arg |  | PD | NA | NA | NA | NA | NA | NA | NA | NA |
| AMPP D-ID-2 | Female | EUR | *GCH1* | p.Lys224Arg |  | PD | 61-65 | NA | NA | Yes | NA | NA | NA | NA |
| GP2-ID-8 | Male | EUR | *GCH1* | p.Lys224Arg |  | PD | 66-70 | Rigidity and bradykinesia | Yes | Yes | No | Father with PD | No | No |
| GP2-ID-9 | Male | EUR | *GCH1* | p.Lys224Arg | *GBA1*<br>p.Arg296Gln | PD | 31-35 | Aching Shoulder, dragging left leg | Yes | Yes | No | Paternal uncle with PD | Slight OFF dystonia | No |
| AMPP D-ID-3 | Female | EUR | *GCH1* | p.Lys224Arg |  | PD | NA | NA | NA | No | NA | NA | NA | NA |
| AMPP D-ID-4 | Male | EUR | *GCH1* | p.Lys224Arg |  | PD | 66-70 | NA | NA | No | NA | NA | NA | NA |
| AMPP D-ID-5 | Male | EUR | *GCH1* | p.Lys224Arg |  | PD | NA | NA | NA | NA | NA | NA | NA | NA |
| GP2-ID-10 | Female | MDE | *GCH1* | p.His210Leu |  | PD | 56-60 | Tremor | Yes | Yes | No | Brother with PD | No | No |
| AMPP D-ID-6 | Male | EUR | *GCH1* | c.626+1del |  | PD | NA | NA | NA | NA | NA | NA | NA | NA |
| GP2-ID-11 | Male |  | *GCH1* | p.Val204Ile |  | PD | 51-55 | Involuntary jaw and tongue movement, rigidity, dragging right foot | Yes | Yes | No | Father with PD | No | No |
| GP2-ID-12 | Male | EUR | *GCH1* | p.Val204Ile |  | PD | 41-45 | Loss of dexterity | Yes | No | No | Not applicable | No | No |
| GP2-ID-13 | Male | EUR | *GCH1* | p.Val204Ile |  | PD | 36-40 | Action tremor | Yes | No | No | NA | No | No |
| AMPP D-ID-7 | Male | EUR | *GCH1* | p.Gly203Arg |  | PD | 56-60 | NA | NA | No | NA | NA | NA | NA |
| GP2-ID-14 | Female | EUR | *GCH1* | p.Thr186Arg |  | PD | 31-35 | NA | NA | No | NA | NA | NA | NA |
| AMPP D-ID-8 | Male | EUR | *GCH1* | p.Glu84* |  | PD | NA | NA | NA | NA | NA | NA | NA | NA |
| GP2-ID-15 | Female | EAS | *PLA2G6* | p.Asp331Tyr | *LRRK2*<br>p.Gly2385Arg | PD | 26-30 | Right leg slowness | Yes | No | Yes | Consanguineous family | Feet and hand dystonia | Severe depression |
| GP2-ID-16 | Male | EAS | *PTS* | p.Pro87Ser;<br>p.Lys91Arg |  | PD | 41-45 | Right leg dystonia | Yes | NA | NA | NA | Right leg dystonia while walking | No |
| Pathogenic variants in combined myoclonus-dystonia genes |  |  |  |  |  |  |  |  |  |  |  |  |  |  |
| GP2-ID-19 | Female | EUR | *SGCE* | c.825+1G>C |  | PD | 36-40 | NA | NA | No | NA | NA | NA | NA |
| Pathogenic variants in isolated dystonia genes |  |  |  |  |  |  |  |  |  |  |  |  |  |  |
| AMPP D-ID-14 | Female | AJ | *KMT2B* | p.Trp1507* |  | PD | 56-60 | NA | NA | NA | NA | NA | NA | NA |
| GP2-ID-20* | Female | EUR | *THAP1* | p.Lys73Thr |  | PD | 36-40 | Tremor, rigidity | Yes | Yes | No | Mother with PD | Slight OFF dystonia | No |
| GP2-ID-21* | Female | EUR | *THAP1* | p.Lys73Thr | *PRKN*<br>p.Pro132Thrfs*9 (het) | PD | 66-70 | Foot Cramps | Yes | Yes | No | Daughter with PD | Slight OFF dystonia | No |
| GP2-ID-22 | Male | EUR | *THAP1* | p.Asp17Glu |  | PD | 36-40 | NA | NA | Yes | NA | Mother with parkinsonism | NA | No |
| GP2-ID-23 | Female | EAS | *THAP1* | p.Ser4Tyr |  | PD | 36-40 | Gait disturbance | Yes | Yes | No | Brother with PD | No | No |
| AMPP D-ID- | Female | EUR | *TOR1A* | p.Glu303del |  | PD | 51-55 | NA | NA | No | NA | NA | NA | NA |
| 15 |  |  |  |  |  |  |  |  |  |  |  |  |  |  |
| GP2-ID-24 | Male | AJ | TOR1A | p.Glu303del |  | PD | 71-75 | Spatial difficulties | Yes | Yes | Yes | Mother with PD; Daughter and two granddaughters with dystonia | No | Memory loss, visual hallucination, RBD |
| GP2-ID-25 | Female | EUR | TOR1A | p.Arg288Gln | GBA1<br>p.Asn409Ser | PD | 41-45 | Tremor, stiffness in the neck | Yes | Possibly | No | Maternal grandfather with possible PD; Mother with AD | Mild dystonic posture in left arm | no |
| GP2-ID-26 | Male | EAS | VPS16 | p.Asn52Lys |  | PD | 46-50 | Upper limb tremors, bradykinesia | Yes | No | No | Not applicable | Right striatal toe, mild camptocormia and pisa syndrome | No |
| GP2-ID-27 | Male | EAS | VPS16 | p.Asn52Lys |  | PD | 31-35 | Tremor, stiffness, slow movements | Yes# | No | No | Not applicable | Blepharospasm | No |
| GP2-ID-28 | Male | EAS | VPS16 | p.Asn52Lys | GBA1<br>p.Leu483Arg,<br>PINK1<br>p.Leu347Pro<br>(het) | PD | 46-50 | Chest muscle pain, limb tremor | Yes# | No | No | Not applicable | Flexed posture of the left hand, intermittent slight OFF dystonia | No |
| GP2-ID-29 | Female | EAS | VPS16 | p.Asn52Lys |  | PD | 46-50 | Tremor | Yes | No | No | Not applicable | No | No |
| GP2-ID-30 | Female | EAS | VPS16 | p.Asn52Lys | GBA1<br>p.Arg502Cys | PD | 46-50 | Right hand tremor | No | Yes | No | Father with PD | No | No |
| GP2-ID-31 | Male | EAS | VPS16 | p.Asn52Lys |  | PD | 56-60 | Unk | Yes# | Yes | No | Mother and brother with PD | No | No |
| GP2-ID-32 | Female | EAS | VPS16 | p.Asn52Lys | LRRK2<br>p.Arg1628Pro | PD | 31-35 | Body weakness associated with cramps | Yes | No | No | Not applicable | Cervical dystonia | Postural hypotension, frequent falls |
| GP2-ID-33 | Male | EAS | VPS16 | p.Asn52Lys |  | PD | 41-45 | Rest tremor, micrographia, slowness in initiating movement | Yes | No | No | Not applicable | No | No |
| GP2-ID-34 | Male | EUR | VPS16 | p.Gln127* |  | PD | 66-70 | NA | NA | Not Reported | NA | NA | NA | Pathology diagnoses: LBD, CAA, AD |
| GP2- | Female | EUR | VPS16 | c.1332-1G>T | GBA1 RecNcil | PD | 36-40 | Slowness | Yes | No | No | NA | Limb dystonia | No |
| ID-35 |  |  |  |  |  |  |  |  |  |  |  |  | (with ON exaggeration) |  |
| GP2-ID-37 | Female | EAS | VPS16 | p.Ala649Serfs*16 |  | PD | 56-60 | Tremor | Yes# | Yes | No | Paternal aunt with PD; Multiple family members (e.g., father and brother) with tremor | Leg dystonia (probably in OFF-medication state) | No |
| GP2-ID-38 | Male | EUR | VPS16 | c.2375+1G>A |  | PD | 81-85 | NA | NA | Not Reported | NA | NA | NA | Pathology diagnoses: LBD, ARTAG, FTLD, SVD |

Table 2 displays the demographic and clinical characteristics of identified variant carriers. The majority with available data on treatment showed a good response to levodopa (n=25/26, missing for n=19). The median age at PD motor symptom onset or PD diagnosis across all carriers was 47 years (IQR: 38-59 years), ranging from 27 to 83 years. Fourteen carriers showed variable signs of dystonia (information unavailable for n=19); which was believed to be related to motor fluctuations due to dopaminergic medications (OFF dystonia) in a subset. While 17 (42.5%, unknown for n=5) carriers had a positive family history of PD, only two individuals had family members with dystonia (unknown for n=19).

Interestingly, one late-onset PD patient (age at onset >70 years) without dystonia, carrying the pathogenic *TOR1A* p.Glu303del variant, had a mother with PD, but also one daughter and two grandchildren with dystonia (Supplementary Figure 2A). Unfortunately, no additional family members were available for genetic testing.

We also identified a pathogenic *THAP1* variant (p.Lys73Thr) in an individual and her mother (Supplementary Figure 2B), both diagnosed with PD and slight foot OFF-dystonia. Interestingly, the mother’s first motor symptom (age at onset >65 years) were foot cramps, while her daughter showed no additional signs of possible dystonia, and had a younger age at onset (<38 years of age).

In addition to these 45 individuals, we identified ten carriers of single heterozygous pathogenic variants in recessively inherited dystonia genes, including *AOPEP* (n=5), *SPR* (n=4), and *HPCA* (n=1) (Supplementary Table 1). We did not detect any pathogenic/likely pathogenic variants in *ANO3, EIF2AK2, GNAL, PRKRA*, *CP, DNAJC12, GLB1, QDPR, SLC6A3, SLC30A10, SLC39A14, TAF1, TH*, *KCTD17,* and *KCNN2.* However, 613 individuals carried rare variants of uncertain significance across all investigated genes (580 with PD, eight with atypical parkinsonism, and 25 unaffected).

## Discussion

Pathogenic variants in dystonia-linked genes were found in less than 1% of PD patients. Despite being rare, they were enriched in PD patients compared to controls and individuals with atypical parkinsonism, where no such variants were identified.

Not surprisingly and in line with previous studies (10,11), variants in *GCH1* were relatively frequent. *GCH1* is known to cause dystonia and parkinsonism, independently or combined. According to a systematic review, 86% of heterozygous *GCH1* carriers present with dystonia to a variable extent, either isolated or combined dystonia-parkinsonism, while 11% have only parkinsonism. (15) Several case series reported PD patients harboring pathogenic *GCH1* with abnormal DaTscan imaging, indicating nigrostriatal dopaminergic deficits consistent with neurodegenerative parkinsonism (11,16,17). Another study suggested *GCH1* variants may lead to parkinsonism by unmasking subclinical nigral pathology, not by causing nigral neurodegeneration. (18) Additionally, according to the most recent PD genome-wide association studies, common variants were nominated as the likely risk factor in the *GCH1* locus. (19,20) Notably, we identified one recurrent *GCH1* variant (p.Lys224Arg) only present in European-ancestry individuals. Although this variant has a conflicting ClinVar interpretation, it was only found in PD patients and absent from controls in our study, and it was significantly more frequent in European-ancestry PD cases from our study compared to ancestry-matched controls obtained from gnomad (Supplementary Table 2). Further, it was present in two PD patients from the same family (sisters), supporting its pathogenic role. In addition to *GCH1*, variants in genes linked to dystonia-parkinsonism (i.e., *ATP1A3, PLA2G6* and *PTS*) collectively accounted for more than half of the identified carriers overall (24/45, 53%).

Interestingly, 47% (n=21/45) of all identified carriers harboured variants in isolated dystonia genes, where parkinsonism is unexpected. (21,22) Amongst those genes was *VPS16*, with the second largest number of pathogenic variants (n=9) observed in this study. We identified one recurrent *VPS16* variant, exclusively present in East Asian individuals. However, although predicted to be pathogenic on ClinVar and reported as disease-causing in Chinese dystonia patients in the homozygous state (23), the overall pathogenicity evaluation remains partially conflicting since this variant is relatively frequent in East Asians controls. In our study, the frequency in East Asian PD cases was higher compared to ancestry-matched controls obtained from gnomad, though this trend was not statistically significant (Supplementary Table 2). Overall, whether these findings in isolated dystonia genes indicate a causal relationship and contribute to the parkinsonian phenotype (variable clinical expressivity), remains elusive. Another, probably more likely, explanation might be that these are incidental findings reflecting reduced penetrance (for dystonia), a phenomenon well known in several monogenic forms of dystonia, especially in DYT-*THAP1* (penetrance ∼50%) and DYT-*TOR1A* (penetrance ∼30%).

Finally, we identified one carrier of a pathogenic variant in the myoclonus-dystonia gene *SGCE*. Notably, due to maternal imprinting, *SGCE* variants show significantly reduced penetrance when inherited maternally, and the phenotype only develops if the variant is inherited paternally. Unfortunately, no additional family members were available for genetic testing, and we were unable to further investigate the inheritance pattern.

This study has some limitations. The availability of clinical data for samples obtained from AMP-PD was very limited; detailed data on dystonia were not available, thereby not allowing us to perform meaningful genotype-phenotype analyses. Further, family members of identified variant carriers were unavailable for genetic testing within this study; precluding us from performing segregation analyses, which would be helpful in assessing pathogenicity, especially in case of genes with reduced penetrance.

In conclusion, pathogenic variants in dystonia-linked genes were present, albeit rare, among PD cases and absent in individuals with atypical parkinsonism and controls. While misdiagnosis cannot be excluded, available clinical and pathological data support a neurodegenerative disease in most cases. The enrichment in PD patients compared to controls suggest that dystonia gene variants may predispose to PD and that there may be potential biological overlap and shared pathways between dystonia and PD. Importantly, our results, alongside previous evidence from screening studies and GWAS, reinforce *GCH1* as a PD-relevant gene holding clinical implications. Similarly, other genes associated with dystonia-parkinsonism may have clinical relevance for PD patients, whereas variants in isolated or myoclonus-dystonia genes more likely represent incidental findings, potentially due to reduced penetrance. Collectively, our results highlight the need for careful interpretation and counseling in clinical genetics regarding a possible role of pathogenic dystonia gene variants in PD patients.

## Data Availability

Data used in the preparation of this article were obtained from the Global Parkinson’s Genetics Program (GP2; https://gp2.org). Specifically, we used Tier 2 data from GP2 release 8 (DOI: 10.5281/zenodo.13755496). Tier 1 data can be accessed by completing a form on the Accelerating Medicines Partnership in Parkinson’s Disease (AMP-PD) website (https://amp-pd.org/register-for-amp-pd). Tier 2 data access requires approval and a Data Use Agreement signed by your institution. AMP-PD data can be accessed through the AMP-PD website (https://amp-pd.org). Qualified researchers are encouraged to apply for direct access to the data through AMP-PD.

## Acknowledgment

We would like to thank Mirja Thomsen and Johannes Krause for determining the episignatures for *KMT2B* variants. This project was supported by the Global Parkinson’s Genetics Program (GP2; https://gp2.org). GP2 is funded by the Aligning Science Across Parkinson’s (ASAP) initiative and implemented by The Michael J. Fox Foundation for Parkinson’s Research (MJFF). For a complete list of GP2 members see doi.org/10.5281/zenodo.7904831.

Additional whole-genome sequencing data used in the preparation of this article were obtained from the Accelerating Medicine Partnership® (AMP®) Parkinson’s Disease (AMP PD) Knowledge Platform. For up-to-date information on the study, visit https://www.amp-pd.org. The AMP® PD program is a public-private partnership managed by the Foundation for the National Institutes of Health and funded by the National Institute of Neurological Disorders and Stroke (NINDS) in partnership with the Aligning Science Across Parkinson’s (ASAP) initiative; Celgene Corporation, a subsidiary of Bristol-Myers Squibb Company; GlaxoSmithKline plc (GSK); The Michael J. Fox Foundation for Parkinson’s Research; Pfizer Inc.; AbbVie Inc.; Sanofi US Services Inc.; and Verily Life Sciences. ACCELERATING MEDICINES PARTNERSHIP and AMP are registered service marks of the U.S. Department of Health and Human Services. Clinical data used in preparation of this article were obtained from the MJFF-sponsored LRRK2 Cohort Consortium (LCC). For up-to-date information on the study, visit www.michaeljfox.org./lcc. The LRRK2 Cohort Consortium is coordinated and funded by The Michael J. Fox Foundation for Parkinson’s Research. The investigators within the LCC provided data, but did not participate in the analysis or writing of this report. The full list of LCC investigators can be found at www.michaeljfox.org/lccinvestigators. This work was supported in part by the Intramural Research Program of the National Institute on Aging, National Institutes of Health, Department of Health and Human Services, project number ZO1 AG000535 and ZIA AG000949. Data and/or samples used in the preparation of this manuscript were obtained from the National Centre of Excellence in Research on Parkinson’s Disease (NCER-PD).

We would like to thank all participants of the Luxembourg Parkinson’s Study for their important support to our research. Furthermore, we acknowledge the joint effort of the National Centre of Excellence in Research on Parkinson’s Disease (NCER-PD) Consortium members from the partner institutions Luxembourg Centre for Systems Biomedicine, Luxembourg Institute of Health, Centre Hospitalier de Luxembourg, and Laboratoire National de Santé generally contributing to the Luxembourg Parkinson’s Study.

Biospecimens used in the analyses presented in this article were obtained from the Northwestern University Movement Disorders Center (MDC) Biorepository. As such, the investigators within MDC Biorepository contributed to the design and implementation of the MDC Biorepository and/or provided data and collected biospecimens but may not have participated in the analysis or writing of this report. MDC Biorepository investigators include Rizwan Akhtar MD, PhD; Tanya Simuni MD; Dimitri Krainc MD, PhD, Puneet Opal MD, PhD; Joanna Blackburn MD; Monika Szela MHA; and Lisa Kinsley MS, CGC. A gift from the Malkin family generously supported the work of the MDC Biorepository.

## Author contributions

LML, KL, and CK were responsible for the study conceptualization and execution. ZHF was responsible for genetic data processing. LML performed the analyses with support from ZHF and KL. AHT, RNA, RA, RB, MF, JF, SJ, BJ, IJKS, RK, GK, CHL, MTP, SBS, TS, JHS, JMS, YWT, RU, ZKW, LW, RMW, KEZ, HRM, NEM, ZJ, TW and SYL contributed samples of and clinical data on identified variant carriers. LML wrote the first draft of the manuscript. All authors read and approved the final version of the manuscript.

