## Supplementary Material for "Rare but Relevant: Assessing Variants in Dystonia-linked Genes in Parkinson’s Disease"

[1] Laboratory of Neurogenetics, National Institute on Aging, Bethesda, Maryland, USA
[2] Institute of Neurogenetics, University of Luebeck, Luebeck, Germany
[3] German Center for Neurodegenerative Diseases (DZNE), Tübingen, Germany
[4] Global Parkinson's Genetics Program (GP2), Maryland, USA
[5] Division of Neurology, Department of Medicine, Faculty of Medicine, University of Malaya, Kuala Lumpur, Malaysia
[6] Neurological Institute, Tel Aviv Sourasky Medical Center, Tel Aviv, Israel; Department of Neurology, Columbia Irving Medical School, New York, NY, USA
[7] National Institute Mongi Ben Hamida of Neurology, Tunis, Tunisia
[8] Neurology Unit, Department of Systems Medicine, Tor Vergata University of Rome, Rome, Italy
[9] Department of Neurology, Northwestern University Feinberg School of Medicine, Chicago, IL, USA
[10] Department of Clinical and Movement Neurosciences, UCL Queen Square Institute of Neurology, London, UK
[11] Parkinson's Disease Center and Movement Disorders Clinic, Department of Neurology, Baylor College of Medicine, Houston, TX, USA
[12] Queen Square Brain Bank for Neurological Disorders, UCL Queen Square Institute of Neurology, London, WC1N 1PJ, UK
[13] Division of Neuropathology, National Hospital for Neurology and Neurosurgery, University College London NHS Foundation Trust, London, UK, WC1N 3BG
[14] Department of Neurology, Seoul National University Hospital, Seoul, South Korea
[15] Luxembourg Centre for Systems Biomedicine (LCSB), University of Luxembourg, Luxembourg
[16] Luxembourg Institute of Health (LIH), Strassen, Luxembourg and Centre Hospitalier de Luxembourg (CHL), Luxembourg
[17] University Hospital Schleswig-Holstein (UKSH), Campus Kiel, Kiel, Germany
[18] Department of Neurology, National Taiwan University Hospital, Taipei, Taiwan
[19] Centre for Preventive Neurology, Wolfson Institute of Population Health, Queen Mary University of London, London, United Kingdom.
[20] Unidad de Trastornos del Movimiento, Servicio de Neurología y Neurofisiología Clínica, Instituto de Biomedicina de Sevilla, Hospital Universitario Virgen del Rocío/Consejo Superior de Investigaciones Científicas (CSIC)/Universidad de Sevilla, Seville, Spain.
[21] Duncan Neurological Research Institute, Texas Children's Hospital, Houston, TX, USA
[22] Mayo Clinic, Jacksonville, FL, USA
[23] Coalition for Aligning Science, Chevy Chase, Maryland, USA
[24] UCL Movement Disorders Centre, University College London, London, UK
[#] Equal last author contribution

### Supplementary Methods

#### Ethics declaration

This study was approved by all ethics committees or institutional review boards of all participating sites and conducted in accordance with the ethical standards of these institutional and national research committees. Informed consent for study participation was obtained from all participants.

#### Study design and participants

We leveraged data from two different sources. First, we included whole-genome sequencing (WGS) from the Global Parkinson's Genetics Program's (GP2) (1) Data Release 8 (DOI 10.5281/zenodo.13755496). Individual study sites were recruited, and samples and data were collected as previously described. (2,3) Individual-level demographic and clinical data were obtained from participating principal investigators and publicly available databases (e.g., for Coriell samples included in GP2). For the purpose of this study, cohort principal investigators, who submitted samples of identified carriers to GP2, were contacted again for detailed data on dystonia symptoms and family history of dystonia (if additional data were available). Additionally, we incorporated WGS data from the Accelerating Medicines Partnership - Parkinson's Disease (AMP-PD) Release 4. AMP-PD leverages data from participants recruited through multiple studies; detailed information is available on the AMP-PD website (https://amp-pd.org). Genetic and clinical data from participants recruited through AP-PD were obtained with local institutional and ethical approvals and appropriate written participant consent. Demographic and clinical data on participants included in this study were obtained with an approved data usage agreement through the Terra platform. Unfortunately, independent study sites and cohort investigators could not be contacted for additional data, unavailable on AMP-PD (e.g., detailed information on dystonia or family history of dystonia). After removing overlapping samples from both datasets, the final sample set comprised 15,739 individuals, including 7,852 PD patients, 4,287 with atypical parkinsonism, and 3,600 unaffected individuals.

#### WGS data processing

WGS data was processed as described before. (2) Briefly, all samples were genome sequenced to an average of 30x coverage with 150bp paired-end reads following Illumina's TruSeq PCR-free library preparation protocol. AMP-PD's functional equivalence pipeline (4) was used to produce the sequence alignment against the GRCh38DH reference genome. We used DeepVariant v.1.6.1 (5) (https://github.com/google/deepvariant) to generate the single-sample variant calls, and joint-genotyping was performed using GLnexus v1.4.3 (https://github.com/dnanexus-rnd/GLnexus) with the preset DeepVariant WGS configuration (6). Genotypes were set to be missing after variant quality control defined as genotype quality >=10, read depth >=10, and heterozygous allele balance between 0.2 and 0.8, and retained high-quality variants with a call rate > 0.95 after quality control. Genetic ancestry was determined using GenoTools v1.2.3 (https://github.com/GP2code/GenoTools) using default settings. (7)

#### Pathogenicity evaluation

The pathogenicity of variants was evaluated using Franklin (https://franklin.genoox.com - Franklin by Genoox) and Varsome (https://varsome.com/) (8), both of which are based on the consensus recommendations of the American College of Medical Genetics and Genomics (ACMG) (9) criteria, MDSGene (https://www.mdsgene.org/methods) and Clinvar (https://www.ncbi.nlm.nih.gov/clinvar/) (10). The evaluation followed a step-wise approach, details are provided in Supplementary Figure 1.

#### *KMT2B* methylation analysis and episignatures

In order to evaluate the pathogenicity of identified variants in the *KMT2B* gene, we performed methylation analyses and determined the specific episignatures for *KMT2B* variants of uncertain significance (VUS), for which enough DNA was available. The analysis was performed as previously described. (11,12)

### Supplementary Results

#### *KMT2B* methylation analysis and episignatures

We performed methylation analyses and determined the specific episignatures for 20 different *KMT2B* VUS. None of the investigated variants showed a methylation profile suggestive of a pathogenic or likely pathogenic role. In contrast, based on these results, these variants should be classified as benign. The detailed results are summarized in Supplementary Table 3.

### Supplementary Figures

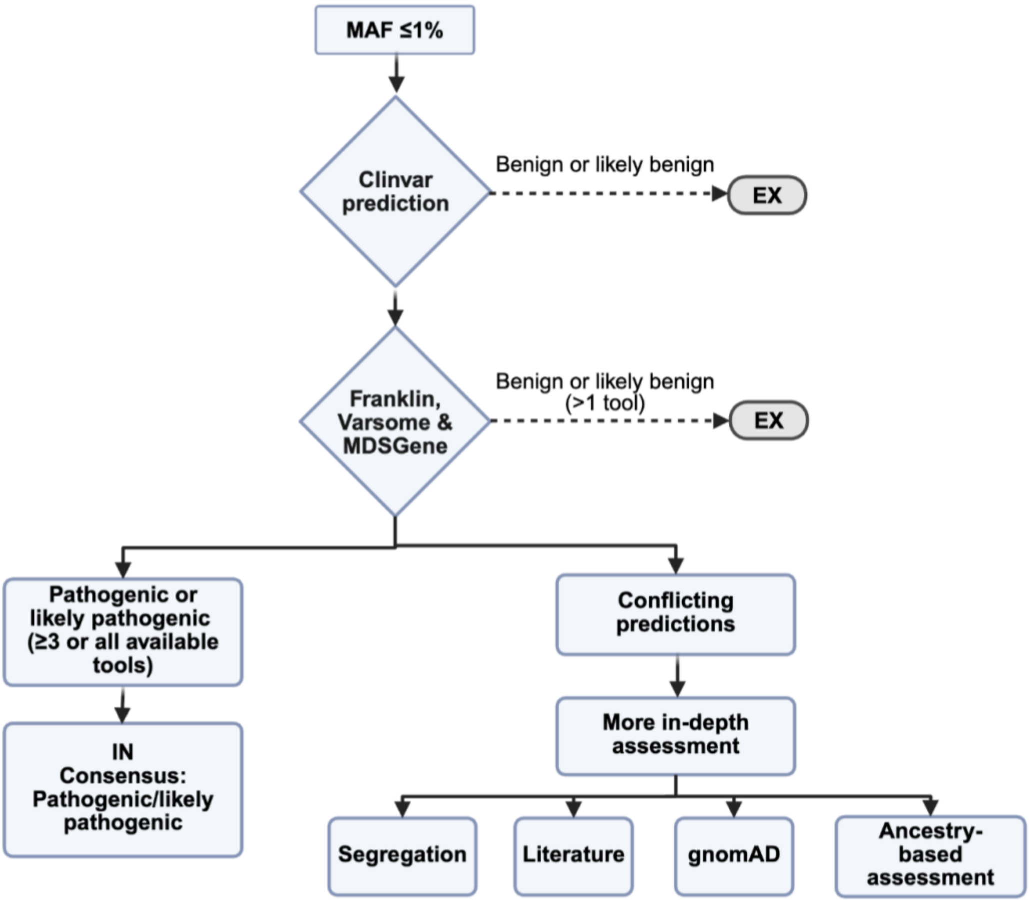

**Supplementary Figure 1. Assessment of pathogenicity.** This flowchart shows the stepwise approach of assessing the pathogenicity of identified variants. First, only variants with a minor allele frequency (MAF) ≤1% based on the overall MAF provided on gnomAD version 4 were extracted from the WGS data. As a next step, all variants predicted to be benign or likely benign on Clinvar were excluded. All remaining variants were evaluated with Franklin, Varsome, and MDSGene. If one out of these tools predicted the variant as benign or likely benign, it was excluded. If, in total, at least three out of four or all available tools predicted the variant to be pathogenic or likely pathogenic, we included the variant. If the predictions were more conflicting, we added another step of a more in-depth analysis. This included i) a segregation analysis (if possible), ii) a brief literature review to identify additional carriers (not captured by MDSGene) or additional evidence from case-control-studies, iii) another more in-depth look into the number of carriers on gnomAD, and iv) an ancestry-specific assessment (e.g., frequency of a given variant in a particular ancestry on gnomAD).

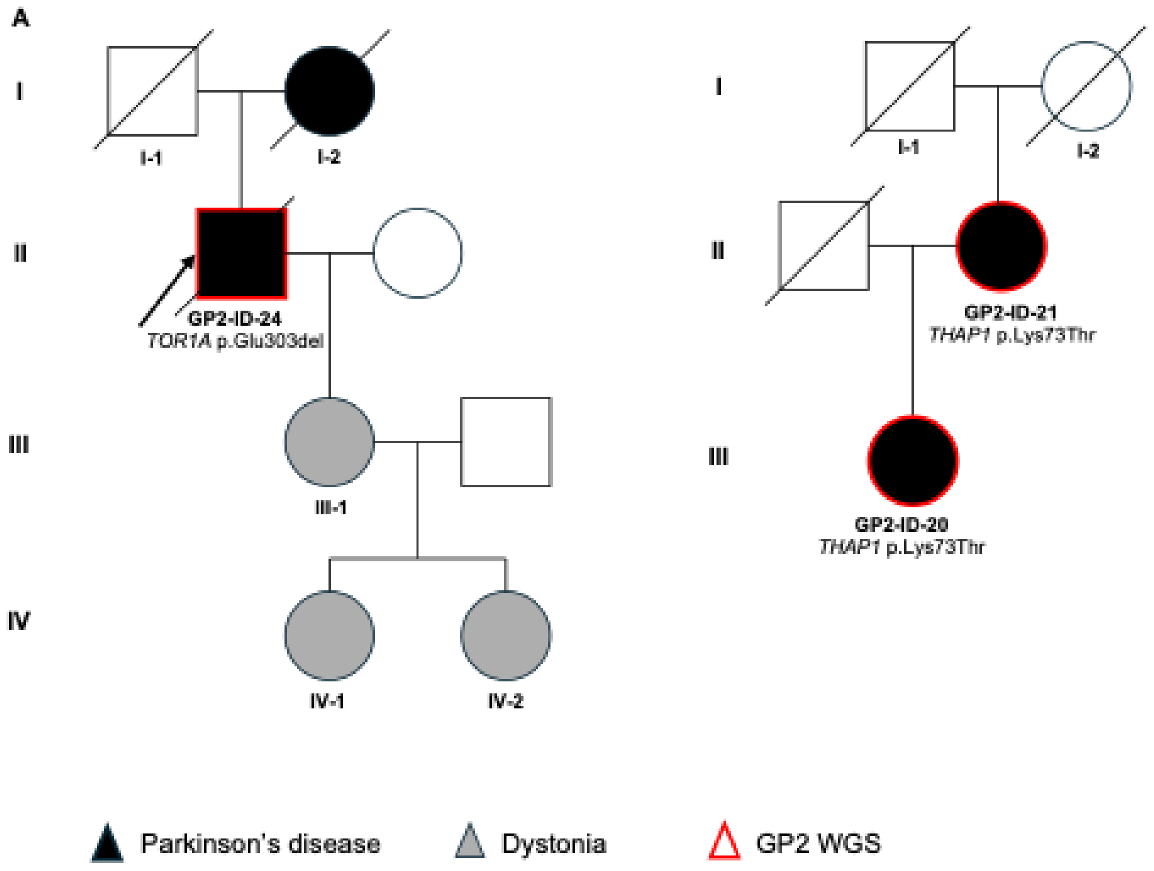

**Supplementary Figure 2. Pedigrees of selected families with individuals harboring pathogenic variants in dystonia-linked genes.** The pedigrees were drawn based on the reported family history and may be incomplete. Squares indicate males, circles indicate females, diamonds indicate unknown sex. Filled symbols indicate affected individuals with Parkinson's disease (PD; black) or dystonia (grey); unfilled symbols indicate unaffected individuals (without PD or dystonia). Individuals highlighted by a red frame underwent short-read whole genome sequencing (WGS) within GP2. (A) Pedigree of individual GP2-ID-24, harboring a pathogenic *TOR1A* p.Glu303del variant. The individual had four daughters, one of which (individual III-1) was diagnosed with dystonia (details unknown) at age 10 years. This individual (III-1) had two daughters (individuals IV-1 and IV-2) with dystonia (details unknown). (B) Pedigree of individuals GP2-ID-20 and GP2-ID-21, both harboring the pathogenic *THAP1* p.Lys73Thr variant.

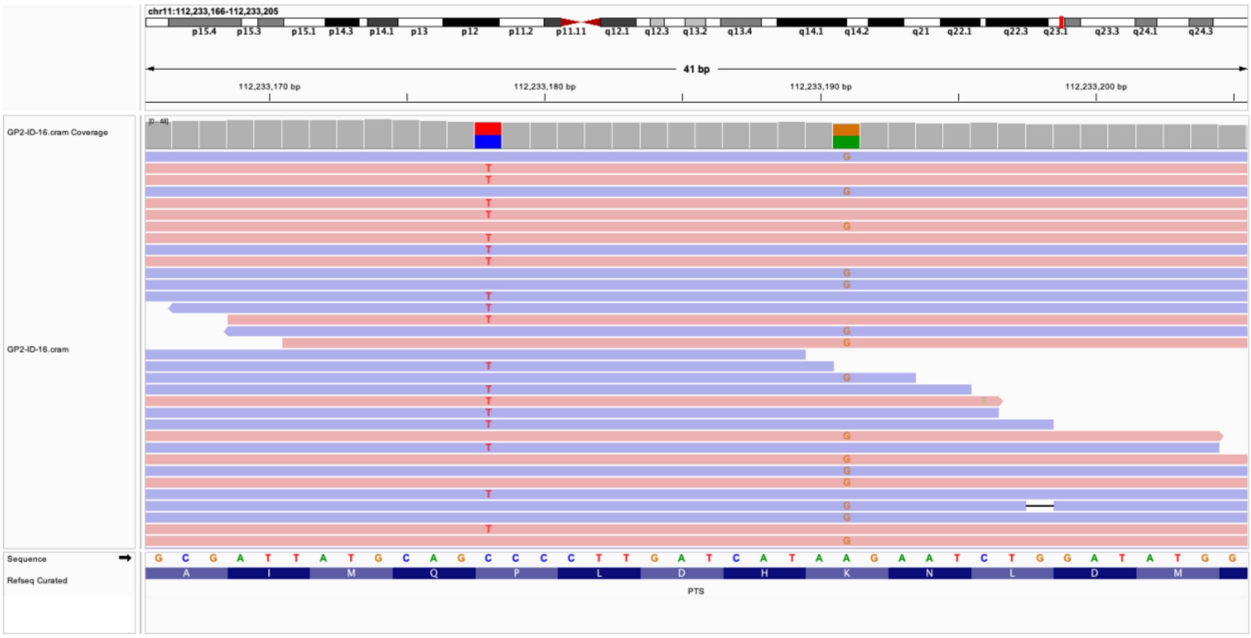

**Supplementary Figure 3. Integrative Genomics Viewer (IGV) screenshot showing compound heterozygous variants in *PTS* in sample GP2-ID-16.** The two heterozygous variants, chr11:112233178:C:T (p.Pro87Ser) and chr11:112233191:A:G (p.Lys91Arg), are located on separate alleles, as indicated by their distribution across distinct sequencing reads. Read depth and variant allele frequencies are visualized across the relevant genomic region, supporting a compound heterozygous configuration in the affected individual.

### Supplementary Tables

#### Supplementary Table 1. Identified carriers of single heterozygous pathogenic variants in recessively inherited dystonia-linked genes.

| | AMPPD-ID-16 | GP2-ID-39 | AMPPD-ID-17 | GP2-ID-40 | GP2-ID-41 | GP2-ID-42 | AMPPD-ID-9 | AMPPD-ID-10 | GP2-ID-17 | GP2-ID-18 |
|---|---|---|---|---|---|---|---|---|---|---|
| **Gender** | Male | Male | Female | Female | Female | Female | ***Male*** | Female | Female | Female |
| **Genetic ancestry** | EUR | EUR | EUR | EUR | EAS | EUR | EUR | EUR | EUR | EUR |
| **Diagnosis** | PD | PD | PD | PD | PD | PD | PD | PD | PD | PD |
| **AAO/AAD** | 56-60 | 56-60 | 46-50 | 36-40 | 41-45 | 71-75 | 56-60 | 51-55 | 31-35 | 66-70 |
| **FH of PD** | Yes | Not reported | Yes | Yes | No | Yes | No | Yes | No | Yes |
| **Gene** | *HPCA* | *AOPEP* | *AOPEP* | *AOPEP* | *AOPEP* | *AOPEP* | SPR | SPR | SPR | SPR |
| **Chromosomal position** | chr1:32889151:T:C | chr9:94760401:C:G | chr9:94800852:CT:C | chr9:94800852:CT:C | chr9:94924098:C:T | chr9:95005584:A:T | chr2:72888457:A:G | chr2:72891406:C:T | chr2:72891466:C:T | chr2:72891502:A:T |
| **Amino acid change** | p.Phe85Leu | p.Tyr206* | p.Val406Cysfs*14 | p.Val406Cysfs*14 | p.Arg493* | p.Lys695* | p.Arg150Gly | p.Arg219* | p.Gln239* | p.Lys251* |
| **Variant type** | missense | nonsense | frameshift | frameshift | nonsense | nonsense | missense | nonsense | nonsense | nonsense |
| **Additional genetic finding** | | | | *GBA1* p.E365K | | | | | | |
| **Pathogenicity consensus** | P/LP | P/LP | P/LP | P/LP | P/LP | P/LP | P/LP | P/LP | P/LP | P/LP |
| **Franklin** | Likely pathogenic | Likely pathogenic | Pathogenic | Pathogenic | Pathogenic | Likely pathogenic | Pathogenic | Pathogenic | Likely pathogenic | Pathogenic |
| **Varsome** | Likely pathogenic | Likely pathogenic | Pathogenic | Pathogenic | Likely pathogenic | Likely pathogenic | Likely pathogenic | Pathogenic | Likely pathogenic | Likely pathogenic |
| **MDSGene** | NA | NA | NA | NA | NA | NA | Definitely pathogenic | Probably pathogenic | NA | Probably pathogenic |
| **ClinVar** | NA | NA | Likely pathogenic | Likely pathogenic | Pathogenic | NA | Pathogenic | Pathogenic | Pathogenic | Pathogenic |
| **CADD** | 29.3 | 24.9 | 2.047 | 2.047 | 38 | 43 | 26.7 | 34 | 37 | 34 |
| **GnomAD AF (total)** | NA | 0.00004709 (total) | 0.00006009 (total) | 0.00006009 (total) | NA | NA | 0.0000591351 (total) | 0.0000197081 (total) | 0.000004336 (total) | 0.0001406 (total) |
| **GnomAD AF (ancestry)** | NA | 0.00006186 (EUR) | 0.00007796 (EUR) | 0.00007796 (EUR) | NA | NA | 0.00009576 (EUR) | 0.000005932 (EUR) | 0.000005085 (EUR) | 0.0001788 (EUR) |

AAO = age at onset, AAD = age at diagnosis, EAS = East Asian, EUR = European, FH = family history, NA = not available/not applicable, EUR = European (non-Finnish), PD = Parkinson's disease, P/LP = pathogenic/likely pathogenic; CADD: Combined Annotation Dependent Depletion; https://cadd.gs.washington.edu/; ClinVar: https://www.ncbi.nlm.nih.gov/clinvar/; GnomAD: https://gnomad.broadinstitute.org/ (version 4.1.0); Franklin: https://franklin.genoox.com; MDSGene: https://www.mdsgene.org/; Varsome: https://varsome.com/

**Supplementary Table 2. Allele frequencies of identified recurrent variants in *GCH1* and *VPS16*.**

| Gene | Variant | Enriched in Ancestry | This study | | | GnomAD | | p value |
|---|---|---|---|---|---|---|---|---|
| ***GCH1*** | chr14:54844099:T:C (p.Lys224Arg) | EUR | **Total AF PD cases (AC)** | **AF EUR PD cases (AC)** | **AF EUR controls (AC)** | **AF total (AC)** | **AF EUR (AC)** | |
| | | | 0.0006368 (10/15704) | 0.0009205 (10/10864) | 0 (0/6096) | 0.0003754 (606/1614128) | 0.0004373 (516/1179952) | **0.325** |
| ***VPS16*** | chr20:2860067:C:A (p.Asn52Lys) | EAS | **Total AF PD cases (AC)** | **AF EAS PD cases (AC)** | **AF EAS controls (AC)** | **AF total (AC)** | **AF EAS (AC)** | |
| | | | 0.0005094 (8/15704) | 0.0032707 (8/2446) | 0 (0/40) | 0.00007435 (120/1614010) | 0.001537 (69/44884) | 0.0619 |

The p value was determined by performing Fisher's exact test comparing the ancestry specific allele frequency in cases from our study with the ancestry specific allele frequency in controls obtained from Gnomad. Statistically significant results are highlighted in bold.
AF = allele frequency, AC = allele count, EAS = East Asian, EUR = European; GnomAD: https://gnomad.broadinstitute.org/ (version 4.1.0)

**Supplementary Table 3. Methylation analyses and episignatures in selected *KMT2B* variants of uncertain significance.**

| *KMT2B* variant | Mean | CV | Interpretation |
|---|---|---|---|
| c.2627_2629del, p.Ala876del | -0.35 | 0.43 | benign |
| c.2726C>A, p.Thr909Lys | -0.61 | 0.35 | benign |
| c.2834T>C, p.Leu945Pro | -0.44 | 0.41 | benign |
| c.3077C>T, p.Thr1026Met | 0.26 | 0.24 | benign |
| c.3109G>A, p.Glu1037Lys | 0.02 | 0.02 | benign |
| c.3191C>T, p.Pro1064Leu | -0.24 | 0.24 | benign |
| c.3868C>T, p.Arg1290Cys | 0.24 | 0.23 | benign |
| c.4072G>T, p.Ala1358Ser | 0.27 | 0.22 | benign |
| c.6106C>T, p.His2036Tyr | 0.19 | 0.16 | benign |
| c.6236G>A, p.Arg2079Gln | -0.18 | 0.14 | benign |
| c.6268G>A, p.Val2090Ile | 0.15 | 0.15 | benign |
| c.6398A>G, p.Glu2133Gly | 0.54 | 0.61 | benign |
| c.5898_5900del, p.Pro1970del | -0.32 | 0.22 | benign |
| c.4803C>G, p.Ile1601Met | 0.94 | 0.64 | benign |
| c.5711C>A, p.Pro1904Gln | -0.86 | 0.63 | benign |
| c.6293C>T, p.Ala2098Val | -0.11 | 0.05 | benign |
| c.7399A>G, p.Ile2467Val | 0.76 | 0.55 | benign |
| c.6664C>G, p.Pro2222Ala | 0.18 | 0.12 | benign |
| c.7008C>G, p.Asp2336Glu | 0.21 | 0.06 | benign |
| c.6944G>A, p.Gly2315Glu | 0.12 | 0.11 | benign |

For each analyzed variant, the mean normalized methylation (Mean) and coefficient of variation (CV) across 113 KMT2B-episignature CpG sites are shown, along with an overall evaluation. Mean values were calculated as the average z-score of methylation deviation from control levels, and CV reflects the homogeneity of this deviation (CV = SD/Mean). The analyses were performed as described. (11,12)
